# A flexible method for optimising sharing of healthcare resources and demand in the context of the COVID-19 pandemic

**DOI:** 10.1101/2020.03.31.20049239

**Authors:** Lucas Lacasa, Robert Challen, Ellen Brooks-Pollock, Leon Danon

## Abstract

As the number of cases of COVID-19 continues to grow, local health services are at risk of being overwhelmed with patients requiring intensive care. We develop and implement an algorithm to provide optimal re-routing strategies to either transfer patients requiring Intensive Care Units (ICU) or ventilators, constrained by feasibility of transfer. We validate our approach with realistic data from the United Kingdom and Spain. In the UK, we consider the National Health Service at the level of trusts and define a 4-regular geometric graph which indicates the four nearest neighbours of any given trust. In Spain we coarse-grain the healthcare system at the level of autonomous communities, and extract similar contact networks. Through random search optimisation we identify the best load sharing strategy, where the cost function to minimise is based on the total number of ICU units above capacity. Our framework is general and flexible allowing for additional criteria, alternative cost functions, and can be extended to other resources beyond ICU units or ventilators. Assuming a uniform ICU demand, we show that it is possible to enable access to ICU for up to 1000 additional cases in the UK in a single step of the algorithm. Under a more realistic and heterogeneous demand, our method is able to balance about 600 beds per step in the Spanish system only using local sharing, and over 1300 using countrywide sharing, potentially saving a large percentage of these lives that would otherwise not have access to ICU.

## 1 Background

The outbreak of COVID-19 [1], the disease caused by the novel coronavirus SARS-CoV-2, detected in China in December 2019 [2], has become pandemic and continues putting national health systems of different countries into significant levels of stress [3, 4, 5, 6] (see [7] and references therein for a detailed overview). Either during the first or successive epidemic waves, the intensive care unit (ICU) demand of several hospitals might surpass their nominal capacity in particular regions in several countries, as has already happened in Italy or Spain [8]. The shortage of sanitary resources is unlikely to be limited to ICU units or ventilators, and other resources will face similar challenges, either during the first surge or in subsequent waves.

In the COVID-19 pandemic, demand for intensive care is not uniform across a country. Epidemic outbreaks can take place in different parts of a country and this can lead to substantial variations of demand both through space and time. Some hospitals may receive substantial numbers of patients early in an outbreak, whilst others may be only mildly affected. This demand heterogeneity opens the possibility of balancing the load of patient admissions such that excessive demand is re-routed to the places which have spare capacity. The clinical need for such a system was evidenced by a spontaneous initiative that took place in Madrid (Spain) in early April 2020 [9], when the Spanish capital was suffering a significant surge of COVID-19 cases. The intensive care lead of 76 hospitals in Madrid created an informal WhatsApp group to share daily information on the ICU demand and availability, with the goal of transferring patients across hospitals in the hope that the network could provide adequate treatment to all patients.

Other tactical load balancing actions have been recently proposed in the US [10, 11]. Of course, this is an example of a quick, crisis emergency action, but as soon as multiple centres are overwhelmed, the demand pattern becomes very complex, and the number of possible transfer combinations increases exponentially. Without a principled and organic approach to patient transfer it is possible to end up worsening the situation. A natural question is thus, given the available resources of a national health system covering a specific region, whether there exist a principled, adaptive and *optimal* way of balancing the demand across hospitals by which a maximal number of patients can receive adequate treatment even during a pronounced epidemic peak, thereby relieving the stress of the whole system. Furthermore, the need to match intensive care supply to patient demand in different parts of the world is indeed currently urgent in areas of the world experiencing serious outbreaks. Here we address such questions by designing and implementing a simple and flexible load sharing procedure which can help to alleviate the level of stress that healthcare systems experience in a systematic way.

The methodology is in principle tailored to address the COVID-19 pandemic situation, but otherwise is general and thus applicable in different countries, at different resolution levels, and for any resource constrained clinical service. The method uses graph-embedded load balancing technology coupled with a simple optimisation kernel, and we showcase its usability by testing it on the UK National Health Service (NHS) and the Spanish health system as examples with different spatial granularity.

Note that graph-embedded load balancing [12,13] has been mainly explored in Computer Science (CS), usually taking a ‘vertex perspective’ for graphical computation with the aim of achieving a centralised solution to load allocation, subject to locality and availability constraints [14]. Interestingly, this line usually relates to minimise large-scale computational efforts, rather than actually sharing physical resources. A similar approach overlaps with the so-called Social Choice Theory of allocating goods among a set of agents under some constraints that overlaps economics, social sciences and computer science [15, 16, 17, 18]. More closely related to our approach is the concept of dynamic load balancing, theoretically explored in the CS literature recently [19, 20]. Similar approaches have also been investigated in the Operations Research (OR) literature, and in particular the topic of location theory is relevant here as well [21, 22]. All these provide a reasonably mature mathematical framework which we subsequently rely on. Indeed, here we build on conceptually similar approaches although we focus on a healthcare network where resources to be shared consist of ICU beds or ventilators, within the context of the COVID-19 pandemic. After presenting the algorithmic modelling, as a proof of concept we apply our framework to two realistic cases at different spatial resolutions: the United Kingdom’s full NHS trust network, and the Spanish contact network between autonomous communities. We focus on the problem of ICU demand, propose and implement a routine strategy to transfer resources across the network, and demonstrate that it is capable of useful and relevant outcomes.

## 2 Materials and Methods

### 2.1 Transfer Networks

We first define a network over which load and resources can be shared. Demand and capacity data –and thus, load sharing–can be coarse-grained at different resolutions: hospitals, postcodes, trusts, and broader regions. In this paper, we consider two levels of resolution: NHS trusts (UK) and autonomous communities (Spain).

#### 2.1.1 NHS trust network

We coarse-grain data for the UK at the level of trusts, as the main units of NHS organisation. We have *N =* 141 trusts across the UK, where each trust corresponds to a conglomerate of hospitals. For each trust, we define a single central position by finding the centroid of the polygon whose vertices are the hospitals belonging to that trust. While spatial coordinates are given in terms of latitude and longitude, we make a small angle approximation and interpret latitude and longitude as cartesian coordinates. In particular, under this approximation the centroid coordinates of trust ireduces to the arithmetic mean of the coordinates of each hospital in the trust

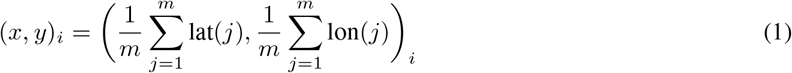

In the event that the net capacity c_i_ of each hospital is also available, then instead of computing the centroid, one can compute the center of mass by appropriately weighting the contribution of each hospital:

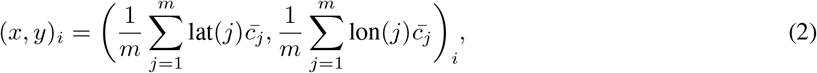

where 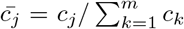 is the normalised capacity of hospital *j*^1^, and m is the number of hospitals in trust *i*. The distance between two trusts corresponds to the Euclidean distance between the centroids or the centers of mass^2^

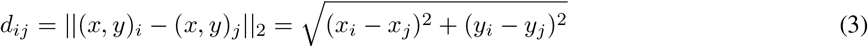

In our case we do not have information on the actual ICU capacity of each specific hospital within a given trust, so we choose to use centroids instead of centers of mass.

Once we have defined the location each of the 141 NHS trusts, we assign a vertex to this spatial location and proceed to tessellate this set. We build a regular geometric graph with degree *k* = 4, where each vertex *i* is connected to the four closest vertices according to the distance *d_ij_* defined above. The resulting graph models the NHS trust network, and each trust will only be allowed to transfer patients or resources to the trusts in their topological neighborhood.

#### 2.1.2 Spain’s autonomous community networks

Spain has a decentralised health system, so we consider that load sharing between hospitals can only take place within each autonomous community (intra-community). Because of that, as a second example here we will consider load sharing at the inter-community level. The network therefore has *N =* 17 nodes, each of them characterising a certain autonomous community. We will consider two different networks: a contact network and a fully connected network. Here, two nodes are connected if the respective autonomous communities share a border. This makes this network more heterogeneous than the NHS trust network, where the maximal degree is *k* = 9 (for the community of *Castilla y Leon* in the centre of Spain). We assume that load sharing can only be performed by road, meaning that this network is disconnected; as two autonomous communities are not part of mainland Spain (Balearic islands and Canary islands). So, we only consider the large connected component, formed by *N* = 15 nodes with varying degree 2 ≤ *k* ≤ 9. Distance is not a constraint in this case.

Additionally, we will also consider a fully connected network formed of *N* = 15 nodes on the mainland, where all possible links are present.. This models the ideal situation where the transfer of ventilators between any two autonomous communities is possible, e.g. using the national train network, as already proposed [23]. The Balearic islands and the Canary islands are, again, not part of this network.

### 2.2 Local load sharing model

The basic architecture of the local load sharing model is depicted in Fig.1. For each node, the algorithm takes `projected-ICU-demand` data (aggregated at the NHS trust level or the autonomous community level, depending on the example), matches with its `baseline-ICU-capacity` (aggregated number of ICU beds or ventilators available), and generates a `local-stress` value for each node. For those nodes where local stress is positive (meaning that demand surpasses capacity), the algorithm explores which neighboring nodes (extracted from the topological neighborhood of the node under analysis) can accept a transfer.

**Figure 1:**
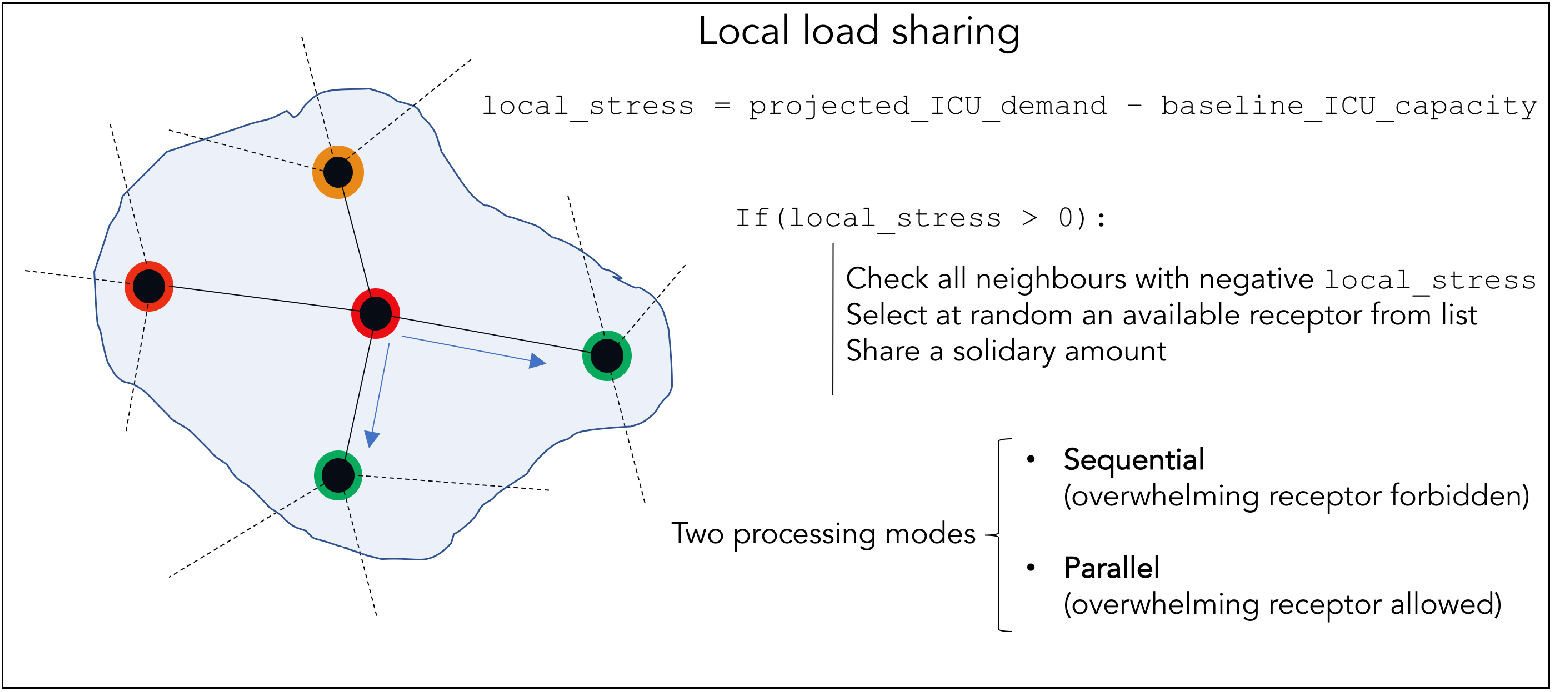
Scheme of the local load sharing model. Red and orange denotes an overwhelmed unit with varying levels of stress, green denotes a unit with capacity.

If more than one receiving node is available in the topological neighborhood of a given node, i.e. there is more than one node with a negative `local-stress` and the distance between origin and destination is smaller than a certain maximally allowed transfer distance *d*_max_, then the algorithm selects the receptor at random. Finally, a solidary load is shared to the receptor. This load is either 50% of the available capacity of the receptor, or the total excess demand of the origin trust, whichever is smaller.

Importantly, the algorithm can perform this analysis either **sequentially** or in **parallel**. In the first case, the `projected-ICU-demand` of each node is sequentially updated after each local load share is performed. This has the positive implication that no receptor can be overwhelmed from the simultaneous load sharing of different nodes. If the update is parallel, overwhelming receiving nodes can occur, but it is also true that more optimal redistributions are available. The implementation asks the user to choose which processing mode is used (sequential or parallel).

### 2.3 Random search optimisation

The basic local load sharing model is run for all nodes (NHS trusts or autonomous communities), and as a result a possible load sharing configuration is extracted, consisting of the specified origin and destination of all the packets of ICU patients shared:

`Trust i shared x loads to trust j`

To assess the global impact of such load sharing configuration, we define the global stress of the whole system

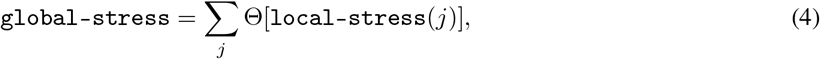

where the sum runs over all trusts *j*, and Θ(*x*) is a rectified linear unit (ReLu), defined by Θ(*x*)= *x* if *x*> 0 and zero otherwise. So essentially `global-stress` counts the total demand of ICU units in excess of capacity, in all those trusts which are projected to be overwhelmed.

Finally, the algorithm is independently run 10^5^ times, and the optimal run with the lowest `global-stress` is retained. In this way, in each realisation the algorithm stochastically chooses a set of actions, and by sampling the search space at random, it finds an approximation to the global load sharing optimum.

### 2.4 Input variables

Now we briefly discuss the main input data required to run the local load sharing model:

- `projected-ICU-demand`: This is an input data to the algorithm and could be estimated following a complex multi-step flow [12], which can be summarised as follows:
  1. The projected number of new infections next week: This quantity can be informed in the first place from an epidemiological model [8, 24] which provides predicted numbers of contagion at different spatial resolutions. Alternatively, or in the absence of such a model, it could be estimated from various sources of data [25] including prescription data [26] or through direct questionnaires^3^. A post-processing of these numbers is then carried out, taking into account (i) age demographics and (ii) associated infection-to-ICU rates.
  2. The projected number of patients already in the hospital which progress to ICU by next week: this number is estimated from real data of hospital admissions and average admission-to ICU likelihood.
  3. The projected number of patients already in ICU this week which will still require ICU next week: this number takes into account both the fatality ratio and the estimated discharge time.
- As a proof of concept, in this work we assume different types of artificial ICU demands (uniform and heterogeneous distributions). We test how the load sharing algorithm performs under different demands.
  - `baseline-ICU-capacity`: This list is extracted from public available databases [27, 28]. In the case of autonomous communities these quantities already have into account some enhancement provided by surge capacity [28], whereas in the case of NHS trusts we only use baseline data, so we expect such capacity to be significantly increased in practice.

## 3 Results

### 3.1 Single-share in the UK NHS trust network

In this first section we assume that each trust can only submit a unique load to a unique receptor trust, to be selected randomly from the trust’s topological neighborhood.

#### 3.1.1 Stress test with fixed, uniform-load ICU demand

As an illustration, we first analyse a stress test case where `projected-ICU-demand` is artificially set to a uniform value of 20 ICU beds per trust (i.e. all trusts receive a demand of 20 beds) whereas we set all `baseline-ICU-capacity` to its real value, and *d*_max_ = ∞. The histogram of `baseline-ICU-capacity` is reported in the top left panel of Fig.2, whereas the histogram of `local-stress`, before and after the load sharing procedure is performed, is depicted in the top right panel of the same figure (we are only showing the parallel mode here). The procedure is capable of reducing the global stress of the system from an initial value of `global-stress` = 611 ICU beds in excess in overwhelmed trusts, to a final value of `global-stress` = 101 after the optimal load sharing is performed, i.e. a transfer and subsequent treatment of 510 ICU patients.

#### 3.1.2 Pipeline of uniform-load stress tests

In a second step, we explore how the system behaves when initial demand per trust varies. To do that, we consider a suite of stress tests and assume for each test that all trusts receive the same load –leading to a uniform demand per trust–, and we compute the `local-stress` before and after the load sharing procedure is applied. Accordingly, the `global-stress` of the whole system and the net reduction in the number of ICU beds in deficit (in collapsed trusts) is also computed.

Results are shown for both the sequential and parallel mode in the bottom panels of Fig. 2. In the bottom left panel we plot the `global-stress` before and after the load sharing procedure is applied, as a function of the initial demand uniformly applied to all trusts. The net reduction (number of ICU patients or ventilators transferred) is plotted in the top right panel. As expected, the `global-stress` curves increase when the demand per trust is increased. At the beginning (for a uniform demand between 0 and 20 ICU beds per trust), the load sharing procedure works very well and completely removes any sign of overwhelming of the system (i.e. keeping the `global-stress` close to zero). When the demand per trust increases further we enter a second regime (between 20 and 40 ICU beds per trust) where the system shows signs of overwhelming but the load sharing procedure still removes a large portion of it (between 40 and 80%). If the demand per trust increases above 40 ICU beds, the whole system becomes overwhelmed, and the load sharing procedure becomes less and less capable of clearing demand, and the resulting net reduction decreases. Results are systematically better for the parallel mode than the sequential mode, but as previously mentioned, this comes at the expense of overwhelming some receptor trusts. Sequential mode still provides very good results and precludes receiving trusts from being overwhelmed.

**Figure 2:**
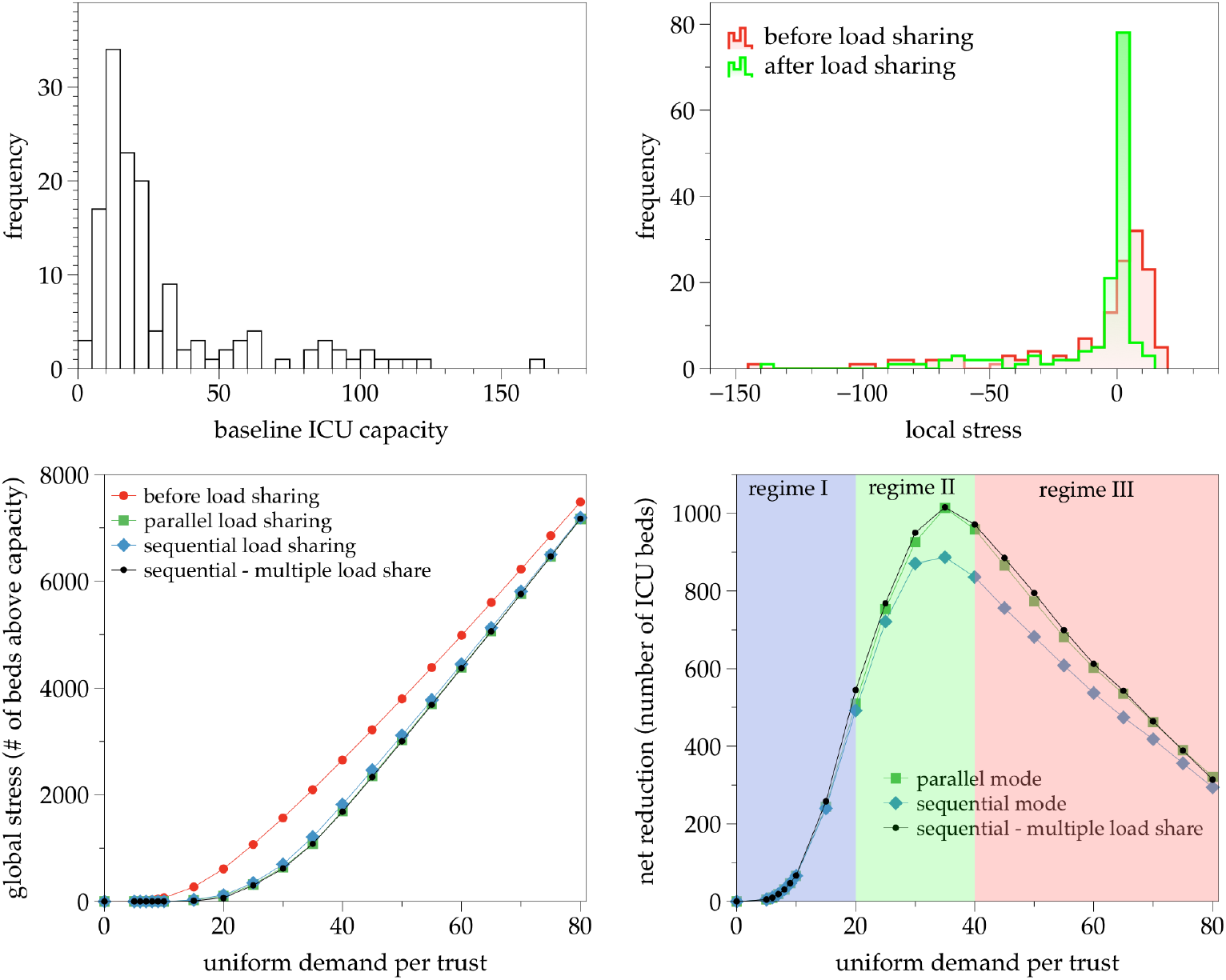
(Top Left panel) Histogram of the `baseline-ICU-capacity` (number of beds) per trust for the NHS trust network. (Top Right panel) Illustration of the histogram of `local-stress` (expected demand of number of beds above capacity) per trust, before and after applying the load sharing procedure. In the synthetic example, all trusts have a uniform `projected-ICU-demand` = 20, whereas the `baseline-ICU-capacity` is informed by data and shown in the left panel. Before the load sharing procedure, `global-stress` = 611, and after the procedure, the new `global-stress` = 101, i.e. a reduction of a total of 510 ICU patients (83%). (Bottom panels) Response of the UK healthcare system in terms of `global-stress` (left) and net reduction in number of ICU beds (right) after the load sharing procedure is applied, as a function of the initial demand per trust (uniform demand across trusts). Different lines correspond to different modes: absence of load sharing (red); single-share, parallel mode (green); single-share, sequential mode (blue); multiple-share, sequential mode (black). Results are similar across modes. We can see three regimes: an initial regime where the load sharing procedure easily removes all signs of overwhelming, a second regime where although the procedure cannot remove all signs of overwhelming, the net reduction is maximised, and a third regime where the load sharing procedure is less and less efficient due to the fact that the whole system is overwhelmed.

### 3.2 Multiple-share in the UK NHS trust network

In this second section we relax the single-share assumption and allow each trust to share multiple loads to multiple receiving trusts, selected from the trust’s topological neighborhood at random. We only consider this option in the ‘sequential processing mode’, where real values of `local-stress` are updated in a sequential way as load sharing is performed.

In the uniform-load stress test, enabling a multiple-share option in the sequential mode provides an improvement in the net reduction of cases when compared to the single-share case. However, the improvement is not large (see Figure 3 for a comparison), and puts the multiple-share sequential mode on a similar footing to the single-share parallel mode, while guaranteeing that no receiving trust is overwhelmed. This result is easy to interpret: there is not much gain in being able to share loads to several receiving nodes (vs one), because on average this possibility will only be useful in a handful of cases. In other words, this result is a byproduct of imposing a uniform-load.

A different result is expected if the initial demand on each node is not uniform. Suppose, for instance, that we have a few trusts that are extremely overwhelmed, and could in principle share loads with several receptors (more than one available receptor in its topological neighborhood), but suppose that those receptors are small trusts with only a small number of available ICU beds. In that case, a single-share approach is clearly deficient, but a multiple-share approach could indeed provide a notable improvement. We illustrate this case in the what follows.

#### 3.2.1 Heterogeneous-load stress test

Instead of loading a uniform demand in each trust, we now test the scenario where demand is heterogeneous, and we only overwhelm ‘large’ trusts. If the trust has a `baseline-ICU-capacity` larger than a certain threshold *τ* then we set an initial value for `projected-ICU-demand` for this trust equivalent to 120% its `baseline-ICU-capacity` (i.e. an increase of 20% above capacity). Similarly, for those trusts whose `baseline-ICU-capacity` is smaller than the threshold *τ* we set an initial `projected-ICU-demand` equivalent to 80% of their corresponding `baseline-ICU-capacity` (i.e. 20% below capacity).

We then apply the load sharing procedure sequentially and compare the net reduction of the global level of stress (number of ICU patients that can be efficiently transferred) for the single-share and the multiple-share options. In Fig. 3 we plot these results as a function of the threshold *τ*, indeed finding that the multiple-share option is much more efficient in this case, as expected.

**Figure 3:**
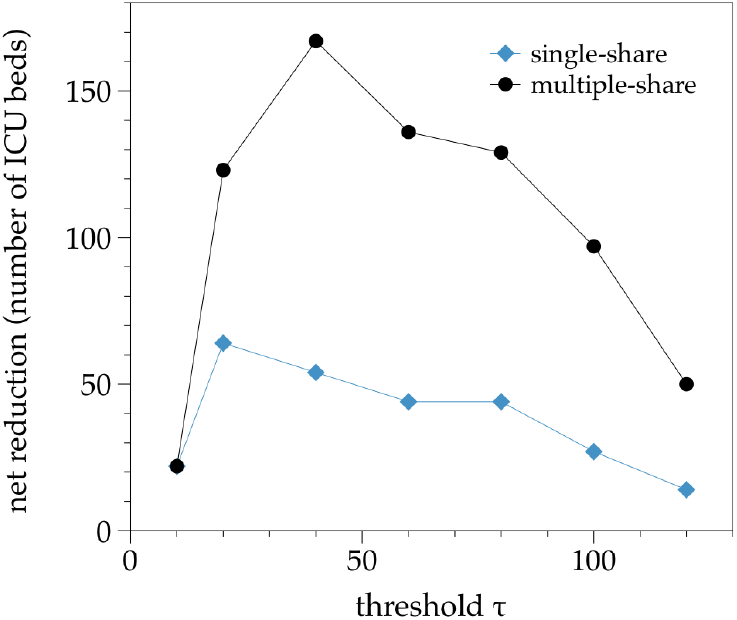
Net reduction offered by the sequential load sharing procedure vs the threshold *τ* (see the text), for a single-share and a multiple-share option, in the UK system undergoing a synthetic heterogeneous-load stress test. The multiple-share option clearly outperforms the single-share one in this case.

### 3.3 Multiple-share in the Spanish autonomous communities contact networks

We now consider the second case: the Spanish healthcare system at the level of spanish autonomous communities. Recall that there are 17 autonomous communities in Spain, and healthcare is decentralised so that each autonomous community runs its own system in a semi-independent way. To explore load sharing effects at the inter-communitary level, instead of adapting the 4-regular network to this context we have constructed two transfer networks: (i) a local contact network of 15 nodes (all autonomous communities in mainland Spain), where two nodes are linked if they share a border, and (ii) a fully connected network of all 15 autonomous communities in mainland Spain.. The former allows for faster transfers, whereas the latter requires using national rail resources [16].

In both cases we use a sequential multiple-share mode. The `ICU-baseline-capacity` for each node is extracted from public data and considers both baseline and surge capacity on 30 March 2020 [22], and the `projected-ICU-demand` is initially set in terms of the ICU occupation number on 30 March 2020 [22]. The average is 63% of the national health system capacity, i.e. all autonomous communities are below capacity showing 63% load. We then increase the demand in each autonomous community, and explore how the load sharing procedure alleviates overwhelming. In the top panels of Fig. 4 we illustrate a scenario, where the Spanish health system is globally overwhelmed (about 200% of the initial demand recorded on the 30th March 2020, or 130% above surge capacity). After load sharing using the contact network, some autonomous communities substantially alleviate such excess and for some others such excess is completely removed. In the ideal scenario where a fully connected network can be used, the load sharing is greatly enhanced.

In the bottom panels of Fig. 4 we plot the global stress and net reduction (total number of ICU beds or ventilators which are effectively transferred) as a function of the national health system saturation (in %), for either using the local contact network or the fully connected network. Both cases enable substantial transfers (about 600 for the local contact network and up to 1300 if transfer is done countrywide, for a single step of the algorithm). Both cases are indeed able to delay global overwhelming, and in the case of the fully connected network the algorithm can maintain the local stress of every autonomous community below capacity even when the true global saturation is around 100%.

For the local contact network, we can distinguish a first phase of steep increase, where only a few communities are overwhelmed and the algorithm is maximally efficient, until the saturation reaches about 120% of the capacity. Then in a second phase, the procedure is still able to transfer many beds or ventilators –even if some autonomous communities will still be overwhelmed–), peaking at a maximum of about 600 beds or ventilators when the system is globally at 170% capacity. As the system gets more and more overwhelmed globally, the load sharing algorithm loses efficiency and the amount of loads that can be shared starts to decrease.

**Figure 4:**
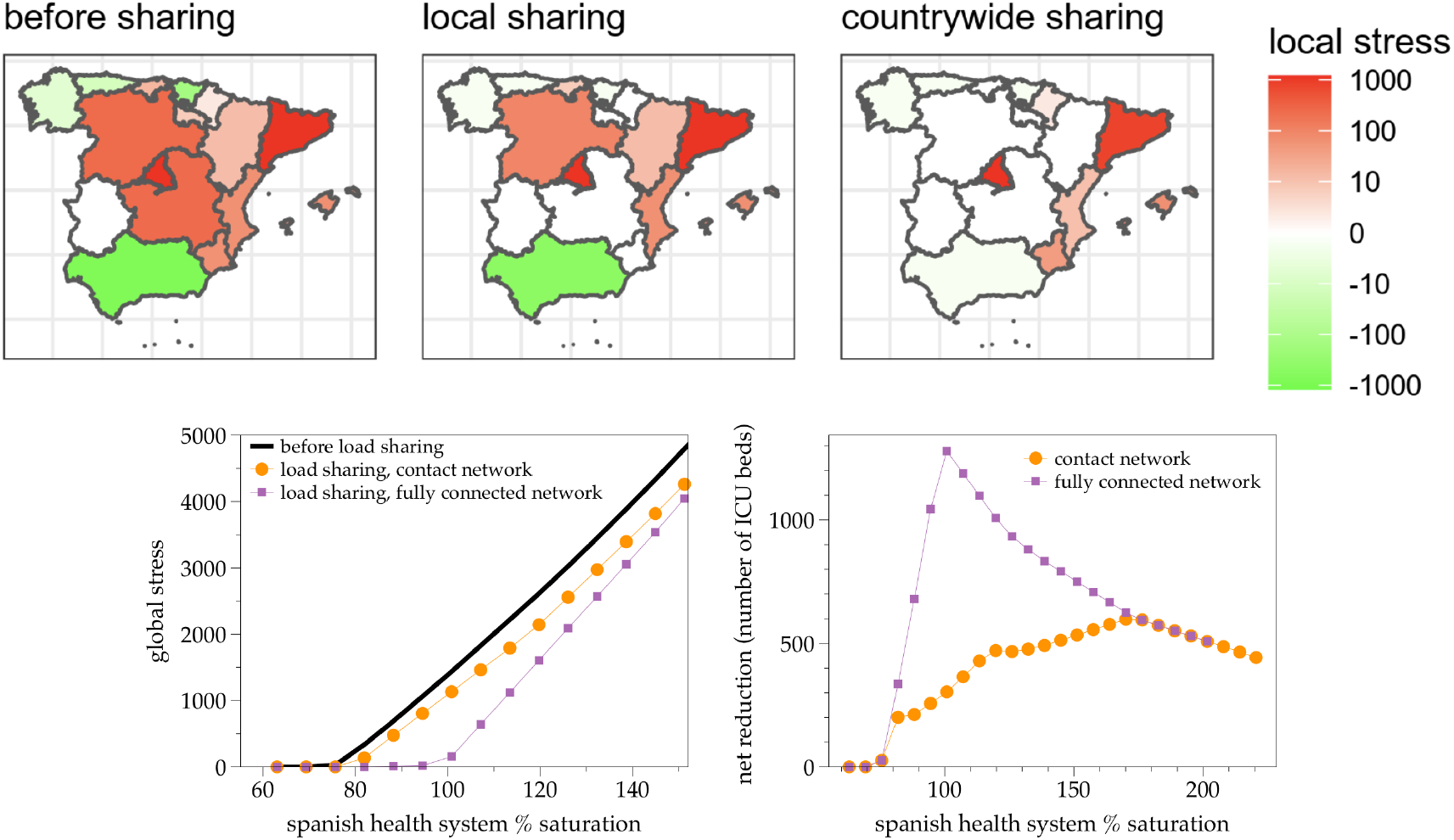
(Top panels) Color-coded local stress of each autonomous community in Spain, before (left panel) and after load sharing (middle and right panels). Canary islands is absent because it is an isolated node and is off-scale. The initial demand is 200% above the real demand as of 30th March 2020, i.e. an approximate demand 130% above surge capacity. In the middle panel a local contact network is used, whereas in the right panel a fully connected network is used. The local contact network is able to transfer and alleviate about 467 ICU beds or ventilators, whereas the fully connected network can transfer 933 units. (Bottom panels) Global stress (left panel) and Net reduction (total number of ICU beds or ventilators efficiently transferred, right panel) offered by the sequential, multiple-share load sharing procedure performed on the Spanish health system, coarse-grained at the level of autonomous communities, as a function of the global saturation percentage of the system (note that capacity has already been enhanced thanks to surge capacity). Orange dots correspond to the contact network, whereas purple squares correspond to the fully connected network.

## 4 Discussion

The COVID-19 pandemic is putting the national health systems of several countries under significant pressure. In this scenario, it is important to devise strategies that distribute capacity of hospitals, not only in terms of the number of ICU beds or ventilators, but also overall capacity (critical care, acute capacity, etc). Here, we have detailed such methodology and have implemented and validated it at two different resolutions: at the level of NHS trusts in the UK and at the level of autonomous communities in Spain. All data and code are available^4^, and will be continually updated. We presented a proof of concept and implementation and showed that this procedure works well and can de-collapse the national health systems in the UK and Spain for a range of scenarios. The random search optimisation layer permits exploration of non-intuitive load sharing configurations which go beyond the simple solution of share load with the neighbor with highest capacity, a strategy which might be locally optimal but also might be leading to a global response far away from the global optimum. We have studied several options, and compared the results of single-share (where a trust can only share load with a single receptor) or multiple-share (where the trust can share parts of the load with different receptors of its neighborhood).

In the context of COVID-19, adopting a load sharing strategy is likely to be beneficial when the whole system is not completely overwhelmed, the projected ICU demand can be accurately estimated, and facilities exist to transfer either patients between ICU departments or ventilators. This is likely early on in the exponential growth phase (of each wave), or in situations where demand is declining either due to interventions or towards the end of the pandemic. When the system is already fully overwhelmed or soon-to-be, this strategy is likely to be inefficient. Furthermore, we also expect this approach to be useful as the epidemic reaches a declining phase, helping to reduce demand and allowing hospitals to return back to normal in a fast an optimised way.

Note that we chose to validate the method in two countries (UK and Spain) as we could focus at two different spatial granularities. However, the method is directly applicable to other countries as well, as long as any sort of transfer system can be put in place. From a clinical point of view, an important point to consider is whether the load sharing can be activated at the ICU stage – potentially leading to transferring highly unstable patients who require ambulance with ICU equipment as well as trained personnel – or if, in anticipation to this, transfer needs to be planned at the point of hospitalisation (admission). In the latter scenario, planning needs to further take into account not only baseline ICU capacity, but overall capacity, also factoring in the estimated lag between admission to hospital and the need for ventilators, which for COVID-19 is currently estimated at about 2 to 3 days. The adequate strategy will also depend on the operational capacity of the system and the country where it is applied to.

For illustration, this work explicitly considers the transfer of ICU patients, however exactly the same approach can be followed if the load to be shared is not patients but ventilators (the units to be moved are not ICU patients but ventilators, so transfer simply happens in the opposite direction, from receptor to origin). Assuming the receptor has both room and personnel to handle additional ventilators, this alternative would indeed (i) eliminate the burden on transferring highly unstable patients and the associated resources required to make such transfers, and (ii) the risk of transferring infection along with patients. Of course, risk (ii) is removed if one only transfers non-COVID ICU patients. In reality, a combination of these mechanisms (transferring ICU patients and ventilators) for sharing load is possible.

This work is subject to several limitations which we hope will be addressed in future work. First of all, the baseline ICU demand only takes into account surge capacity in the Spanish case: more realistic analysis of the UK case shall include surge capacity, that is expected to significantly increase the real ICU capacity of each trust. Second, in the sequential case (where receptors cannot be overwhelmed), overwhelmed nodes can at most share all the excess load, but not more (this latter case would be beneficial if e.g. two-step sharing is needed), therefore multiple-step load sharing strategies have not been explored. Also, the optimisation process implemented here is based on a stochastic search, so there is no rigorous guarantee that the suggested configuration is indeed the global optimum. Other refined methods such as hill climbing, genetic algorithms or simulated annealing could be used to refine this layer, if needed. Other extensions of interest include questions related to dynamic load balancing where the demand varies dynamically Finally, we have assumed that the cost of transfer is zero, i.e. the number of ambulances or the human resources are not a constraint, and that there are enough vehicles to transfer ICU patients or ventilators effectively and enough qualified personnel to handle them. All these limitations can be addressed by suitably extending the specifications of the algorithm, leading to multi-criteria optimisation problems.

## Data Availability

Codes and data are available at
https://github.com/lucaslacasa/loadsharing

## Acknowledgments

LL gratefully acknowledges the financial support of the EPSRC via Early Career Fellowship EP/P01660X/1. RCh gratefully acknowledges the financial support of the EPSRC via grant EP/N014391/1 and NHS England, Global Digital Exemplar programme. LD gratefully acknowledges the financial support of the Medical Research Council and The Alan Turing Institute under the EPSRC grant EP/N510129/1.

## Author contributions

LL, RC and LD conceived research. LL and RC conducted research, curated data, implemented algorithms and performed the data analysis. All authors discussed results and wrote the paper.

## Footnotes

1 we normalise it such that x and y still have dimensions of length

2 if more precise coordinates need to be used, instead of these we can use Haversine formula.

3 Data can be retrieved and processed from apps and other surveillance systems such as centralised webpages where citizens submit their symptoms. These questionnaires, coupled with a classification algorithm, can estimate the number of latent infected people in a certain region or postcode.

4 https://github.com/lucaslacasa/loadsharing

